# Characterization of anti-viral immunity in recovered individuals infected by SARS-CoV-2

**DOI:** 10.1101/2020.03.17.20036640

**Authors:** Ling Ni, Fang Ye, Meng-Li Chen, Yu Feng, Yong-Qiang Deng, Hui Zhao, Peng Wei, Jiwan Ge, Xiaoli Li, Lin Sun, Pengzhi Wang, Peng Liang, Han Guo, Xinquan Wang, Cheng-Feng Qin, Fang Chen, Chen Dong

**Author notes:** To whom correspondence should be addressed: Chen Dong,.cn; or Fang Chen. These authors contributed equally to this work (Ling Ni, Fang Ye, Meng-Li Chen).

## Abstract

The WHO has declared SARS-CoV-2 outbreak a public health emergency of international concern. However, to date, there was hardly any study in characterizing the immune responses, especially adaptive immune responses to SARS-CoV-2 infection. In this study, we collected blood from COVID-19 patients who have recently become virus-free and therefore were discharged, and analyzed their SARS-CoV-2-specific antibody and T cell responses. We observed SARS-CoV-2-specific humoral and cellular immunity in the patients. Both were detected in newly discharged patients, suggesting both participate in immune-mediated protection to viral infection. However, follow-up patients (2 weeks post discharge) exhibited high titers of IgG antibodies, but with low levels of virus-specific T cells, suggesting that they may enter a quiescent state. Our work has thus provided a basis for further analysis of protective immunity to SARS-CoV-2, and understanding the pathogenesis of COVID-19, especially in the severe cases. It has also implications in designing an effective vaccine to protect and treat SARS-CoV-2 infection.

---

At the end of 2019, patients with Coronavirus Disease 2019 (COVID-19) were identified in Wuhan, China ^1^, infected by a novel coronavirus, now named as severe acute respiratory syndrome coronavirus 2, SARS-CoV-2. The WHO then declared this outbreak a public health emergency of international concern ^2^. The genome sequence of SARS-CoV-2 bears 96% ^3^ and 79.5% identity to that of a bat coronavirus and SARS-CoV, respectively ^4^. Like SARS-CoV and MERS-CoV, SARS-CoV-2 belongs to the beta genus Coronavirus in the Corornaviridae family ^5^. Clinically, several papers showed that most COVID-19 patients developed lymphopenia as well as pneumonia with higher plasma levels of pro-inflammatory cytokines in severe cases ^6-8^, suggesting that the host immune system is involved in the pathogenesis ^9,10^. Patients infected by SARS-CoV or MERS-CoV were previously reported to have antibody responses^11-14^, but exhibited defective expression of type I and II interferon (IFN), indicative of poor protective immune responses ^15-17^. However, to date, there was hardly any study in characterizing the immune responses, especially adaptive immune responses to SARS-CoV-2 infection. Only one COVID-19 patient was shown with nucleocapsid protein (NP)-specific antibody response, in which IgM peaked at day 9 after disease onset and then switched to IgG by week 2 ^3^, suggesting involvement of humoral immunity. SARS-CoV-2-specific T lymphocyte response was unclear. In this study, we collected blood from COVID-19 patients who have recently become virus-free and therefore were discharged, and analyzed their SARS-CoV-2-specific antibody and T cell responses.

Clinical and pathological characteristics of the 12 COVID-19 patients in this study were shown in Table 1. All the patients initially showed mild symptoms via CT scan and were positive during SARS-CoV-2 nucleic acid testing. Of them, 6 (patients #1-6) were newly discharged, while the remaining 6 were 2 weeks post discharge (follow-up patients, patients #7-12). In line with the previous reports, 2 patients (#5, 8) showed lymphopenia (normal range is 1.1-3.2X10e9 cells per L). However, only two travelled to Wuhan city within the past 3 months. One serum was obtained before the SARS-CoV-2 outbreak (healthy donor #1). 3 additional healthy donors (#2-4) who had been in close contacts with the patients were recruited in this study. Human AB serum (GemCell, CA) was used as a negative control.

**Table 1.**
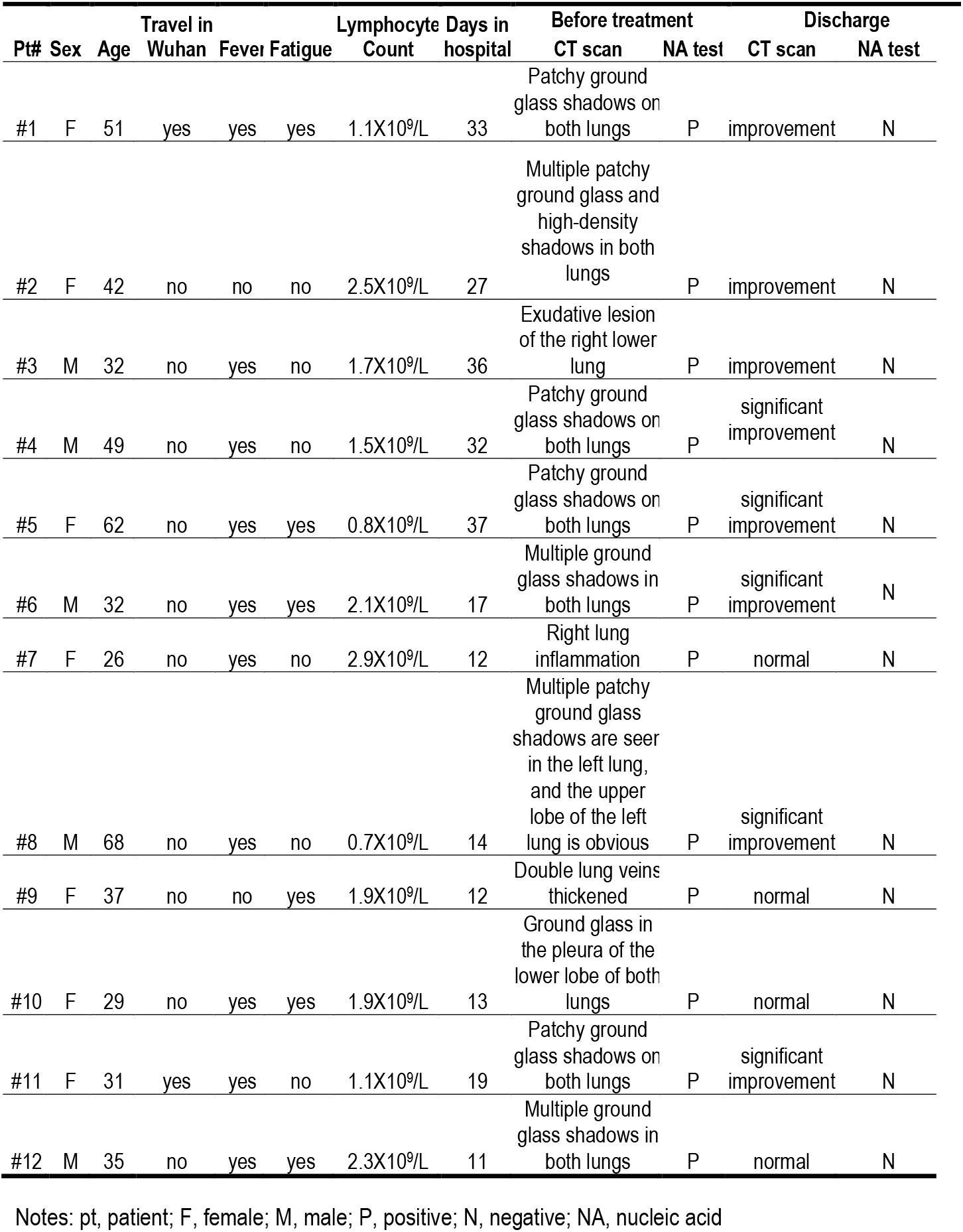
Clinical and pathological characteristics of the COVID-19 patients

| Pt# | Sex | Age | Travel in Wuhan | Fever | Fatigue | Lymphocyte Count | Days in hospital | Before treatment<br>CT scan | NA test | Discharge<br>CT scan | NA test |
| --- | --- | --- | --- | --- | --- | --- | --- | --- | --- | --- | --- |
| #1 | F | 51 | yes | yes | yes | 1.1X10<sup>9</sup>/L | 33 | Patchy ground glass shadows or both lungs | P | improvement | N |
| #2 | F | 42 | no | no | no | 2.5X10<sup>9</sup>/L | 27 | Multiple patchy ground glass and high-density shadows in both lungs | P | improvement | N |
| #3 | M | 32 | no | yes | no | 1.7X10<sup>9</sup>/L | 36 | Exudative lesion of the right lower lung | P | improvement | N |
| #4 | M | 49 | no | yes | no | 1.5X10<sup>9</sup>/L | 32 | Patchy ground glass shadows or both lungs | P | significant improvement | N |
| #5 | F | 62 | no | yes | yes | 0.8X10<sup>9</sup>/L | 37 | Patchy ground glass shadows or both lungs | P | significant improvement | N |
| #6 | M | 32 | no | yes | yes | 2.1X10<sup>9</sup>/L | 17 | Multiple ground glass shadows in both lungs | P | significant improvement | N |
| #7 | F | 26 | no | yes | no | 2.9X10<sup>9</sup>/L | 12 | Right lung inflammation | P | normal | N |
| #8 | M | 68 | no | yes | no | 0.7X10<sup>9</sup>/L | 14 | Multiple patchy ground glass shadows are seen in the left lung, and the upper lobe of the left lung is obvious | P | significant improvement | N |
| #9 | F | 37 | no | no | yes | 1.9X10<sup>9</sup>/L | 12 | Double lung veins thickened | P | normal | N |
| #10 | F | 29 | no | yes | yes | 1.9X10<sup>9</sup>/L | 13 | Ground glass in the pleura of the lower lobe of both lungs | P | normal | N |
| #11 | F | 31 | yes | yes | no | 1.1X10<sup>9</sup>/L | 19 | Patchy ground glass shadows or both lungs | P | significant improvement | N |
| #12 | M | 35 | no | yes | yes | 2.3X10<sup>9</sup>/L | 11 | Multiple ground glass shadows in both lungs | P | normal | N |

Notes: pt, patient; F, female; M, male; P, positive; N, negative; NA, nucleic acid

In order to detect anti-viral immune responses, we first constructed recombinant pET28-N-6XHis by linking 6 copies of His tag to the C-terminus of NP in the pET28-N vector (Biomed, Cat. number: BM2640). Escherichia coli transformed with pET28-N-6xHis was lysed and tested by Coomassie blue staining to confirm NP expression at 45.51 kDa. NP was further purified by Ni-NTA affinity chromatography and gel filtration. The purity of NP was approximately 90% (Figure S1A). The presence of NP was subsequently confirmed by anti-Flag antibody (Figure S1B). The RBD region of S protein (S-RBD) and main protease (doi: https://doi.org/10.1101/2020.02.19.956235) were produced by a baculovirus insect expression system and purified to reach the purity of 90% (Figure S1A).

Using sera from patients and healthy donors, IgG and IgM against SARS-CoV-2 NP, main protease and S-RBD antigens were analyzed. There was no significant antibody response to main protease in sera from several patients (data not shown), suggesting that it may not serve as an antigen for humoral immunity. We thus focused on NP and S-RBD. The serum from a patient and human AB serum were titrated in order to determine optimal dilutions (data not shown). Dilution of 1:50 was used for IgM and 1:150 for IgG. NP- and S-RBD-specific IgM and IgG antibodies were both detected in the sera of newly discharged patients, compared with healthy donor groups (Figure 1A). Anti-SARS-CoV-2 IgG antibodies were also more obviously observed than IgM in the follow-up patients (#7-12), when compared with healthy donors (Figure 1A). Taken together, these findings indicate that COVID-19 patients mounted IgG and IgM responses to SARS-CoV-2 proteins, especially NP and S-RBD, and also suggest that infected patients could maintain their IgG levels, at least for two weeks.

**Figure 1.**
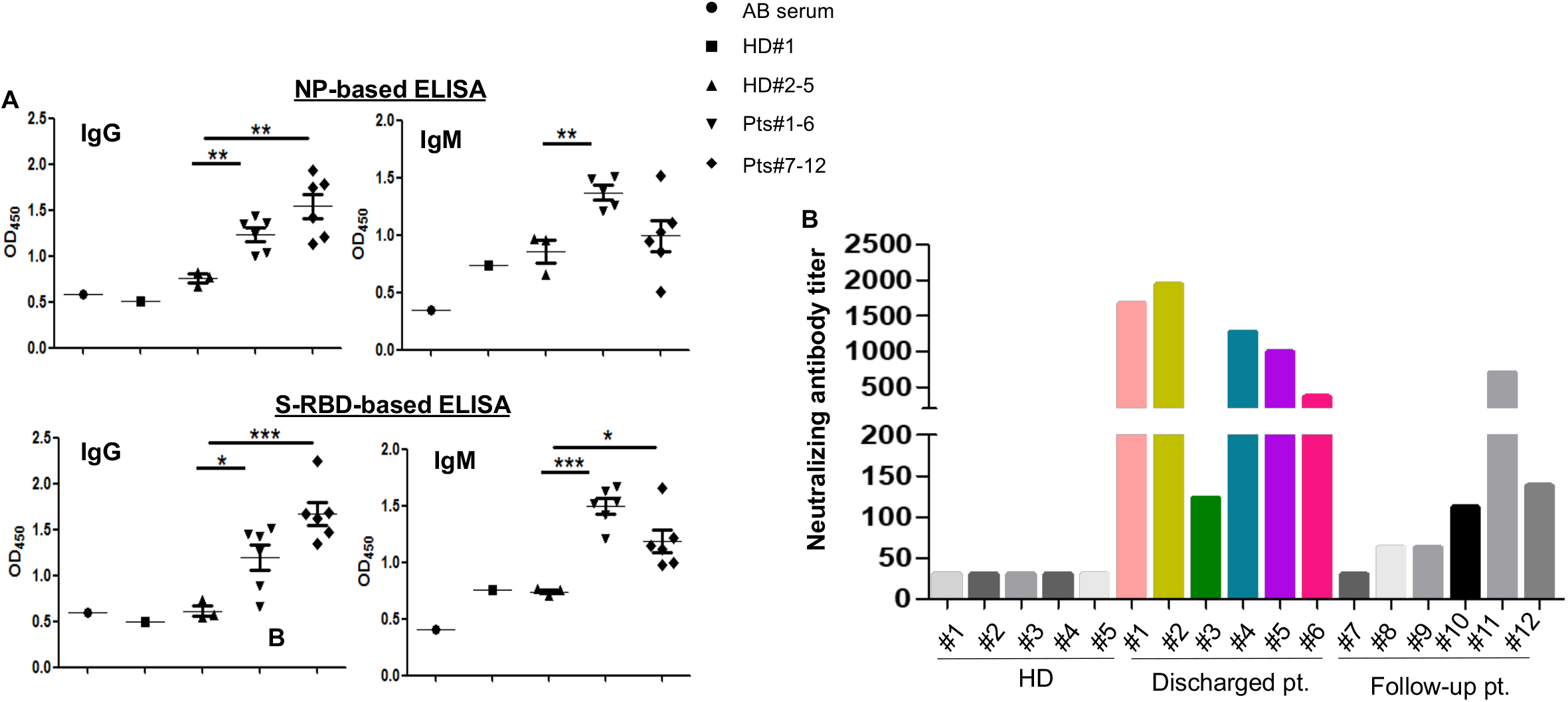
Detection of antibody responses to recombinant SARS-CoV-2 proteins in COVID-19 patients. (A) Serological responses of 12 COVID-19 patients to recombinant NP (top) and S-RBD (bottom). The experiment was performed in duplicates. (B) Measurement of neutralizing antibody titers by pseudovirus-based assay. The experiment was performed in triplicates. NP, nucleocapsid protein. S-RBD, receptor binding domain of spike protein. HD, healthy donor. Pt, patient. HD#1, the serum was collected in 2018. HD#2-4, the sera were from close contacts. *P<0.05, 0.05<**P<0.001, ***P<0.001.

Since RBD domain of the S protein has been shown to bind to human receptor ACE2 ^3^, the existence of antibodies against it may suggest neutralization of SARS-CoV-2 infection. To assess this, we performed pseudovirus particle-based neutralization assay. As shown in Figure 1B, patients #1, 2, 4 and 5, all within the discharged group, had high levels of neutralizing antibody titers. These results demonstrate that most recently discharged patients had protective humoral immunity to SARS-CoV-2. All except patient #11, the follow-up patients had lower levels of neutralizing antibody titers than recently discharged patients, though all positive with the exception of patient #7 being negative. Whether protective humoral immunity can be maintained needs further investigation.

To explore cellular immune responses to SARS-CoV-2, we isolated PBMCs from the whole blood and phenotypically analyzed them by flow cytometry (Figure 2A). We found that compared to discharged patients, there was a trend towards an increased frequency of NK cells in the follow-up patients (Figure S2). However, there was no significant difference in terms of the percentages of T cells among those two groups and the healthy donors.

**Figure 2.**
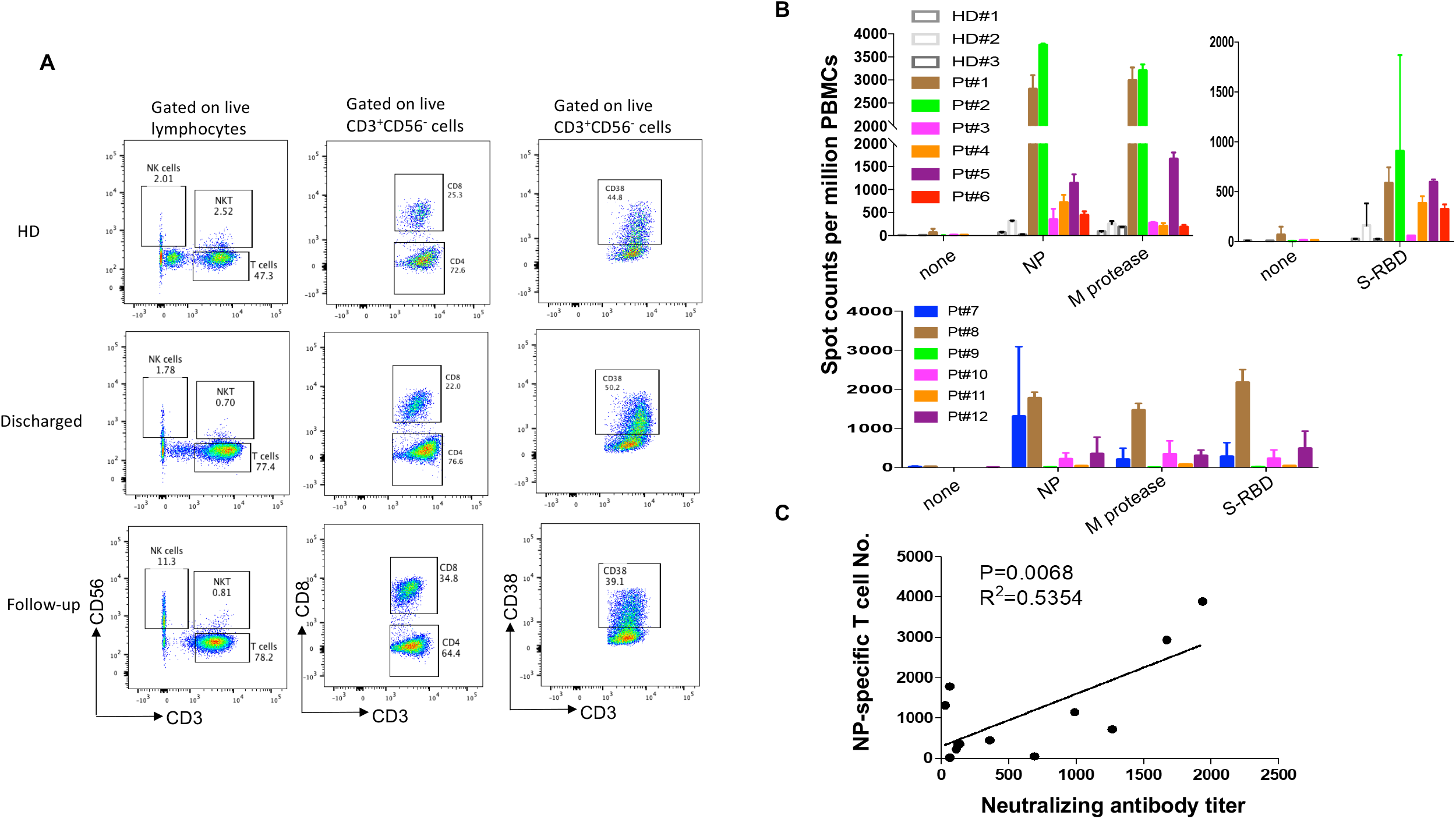
T cell responses to recombinant SARS-CoV-2 proteins in COVID-19 patients. (A) Phenotypic analysis of PBMCs from representative COVID-19 patients. (B) IFN-γ ELISpot analysis of COVID-19 patients to recombinant proteins. The experiment was performed in duplicates. (C) Correlation analysis of the neutralizing antibody titers with the numbers of NP-specific T cells (n=12). M protease, main protease. NP, nucleocapsid protein. S-RBD, receptor binding domain of spike protein.

To assess virus-specific cellular immunity, we then treated PBMCs with recombinant NP, main protease and S-RBD, followed by IFN-γ ELISpot analysis. The results were considered positive if there were at least 1-fold increase in the numbers of IFN-γ-secreting T cells in the subject than in the healthy donors. As shown in Figure 2B, compared with healthy donors, the numbers of IFN-γ-secreting NP-specific T cells in patients #1, 2, 4 and 5 were much higher than other patients, suggesting that they had developed SARS-CoV-2-specific T cell responses. Of note, patients #1, 2, 4 and 5 developed both strong humoral and cellular immune responses. Main protease-specific T cells were detected in patient #1, 2 and 5, while patients # 1, 2, 4, 5 and 6 showed S-RBD-specific T cells. Although the numbers of IFN-γ-secreting S-RBD specific T cells were much lower than those of NP-specific T cells, they could be detected in more patients than those for other viral proteins. S-RBD thus not only elicited humoral immunity that may result in blockade of receptor binding during viral entry in host cells, but also induced T cell immune responses, suggesting S-RBD is a promising target for SARS-CoV-2 vaccines. In the follow-up patients, only patient #8 who showed lymphopenia before treatment still had a high number of IFN-γ-secreting T cells in response to NP, main protease and S-RBD (Figure 2B), which suggests that anti-viral T cells may not be maintained at high numbers in the PBMCs in the recovered patients. More interestingly, when combining all 12 patients in our analysis, there was a significant correlation between the neutralizing antibody titers and the numbers of NP-specific T cells (Figure 2C), indicating that the development of neutralizing antibodies may be correlated with the activation of anti-viral T cells. Thus, effective clearance of virus may need collaborative humoral and cellular immune responses.

In summary, we provided the first analysis of SARS-CoV-2-specific humoral and cellular immunity. Both were detected in newly discharged patients, suggesting both participate in immune-mediated protection to viral infection. However, follow-up patients exhibited high titers of IgG antibodies, but with low levels of virus-specific T cells, suggesting that they may enter a quiescent state.

Our work has thus provided a basis for further analysis of protective immunity to SARS-CoV-2, and understanding the pathogenesis of COVID-19, especially in the severe cases. It has also implications in designing an effective vaccine to protect and treat SARS-CoV-2 infection.

## Materials and Methods Ethics statement

All procedures followed were in accordance with the ethical standards of the responsible committee on human experimentation (the institutional review board at Tsinghua University) and with the Helsinki Declaration of 1975, as revised in 2000. Informed consent was obtained from all subjects for being included in the study. All patient data were anonymized before study inclusion. The blood samples of COVID-19 patients and healthy donors were obtained from Chui Yang Liu Hospital affiliated to Tsinghua University in Beijing.

### Expression and Purification of recombinant proteins

The recombinant His-tagged NP of SARS-CoV-2 was expressed in E. coli by a T7 expression system, with 1 mM IPTG induction at 37 °C for 4 h. pET-28aHis-tagged S-RBD (amino acids 319-541) was expressed by a Baculovirus system in insect cells (doi: https://doi.org/10.1101/2020.02.19.956235). Purified proteins were identified by SDS-PAGE gels and stained with Coomassie blue. Western blot was performed to confirm their antigenicity by mouse anti-His monoclonal antibody (Proteintech, HRP-66005).

### Isolation of PBMC

PBMCs were isolated from anti-coagulant blood using Ficoll-Hypaque gradients (GE Healthcare Life Sciences, Philadelphia, PA) as previously described ^18^ under the BSL-3 facility in AMMS.

### Anti-SARS-CoV-2 IgG/IgM ELISA

For IgM/IgG testing, 96-well ELISA plates were coated overnight with recombinant NP and S-RBD (100 ng/well). The sera from COVID-19 patients were incubated for 1 h at 37°C. An anti-Human IgG-biotin conjugated monoclonal antibody (Sino Biological Inc., Wayne, PA) and streptavidin-HRP were used at a dilution of 1:500 and 1:250, respectively, and anti-human IgM-HRP conjugated monoclonal antibody (Inova Diagnostics, Inc., San Diego, CA). The OD value at 450 was calculated.

### Neutralizing antibody assay

Pseudovirus expressing the SARS-CoV-2 S protein was produced as described previously ^19^. pNL43Luci and GP-pCAGGS were co-transfected into 293T cells. 48 hours later, SARS-CoV-2 pseudovirus-containing supernatants were mixed with at least 6 serially diluted serum samples from the COVID-19 patients at 37°C for 1 hour. Then the mixtures were transferred to 96-well plates containing monolayers of Huh-7 cells. 3 hours later, the medium was replaced. After incubation for 48 h, the cells were washed, harvested in lysis buffer and analyzed for luciferase activity by the addition of luciferase substrate. Inhibition rate = [1-(the sample group-the cell control group) / (the virus control group-the cell control group)] × 100%. The neutralizing antibody titer were calculated by performing S-fit analysis via Graphpad Prism 7 software.

### Interferon Gamma (IFN-γ) ELISpot

IFN-γ-secreting T cells were detected by Human IFN-γ ELISpot^pro^ kits (MABTECH AB, Sweden) according to the manufacture protocol. Fresh PBMCs were plated in duplicate at 150k per well and then incubated 48h with 1uM of recombinant proteins. Spots were then counted using an ELIspot Reader System (AT-Spot2100, atyx). The number of spots was converted into the number of spots per million cells and the mean of duplicate wells plotted.

### FACS staining

PBMCs were washed with PBS plus 2% FBS (Gibco, Grand Island, NY), and then Fc blocking reagent (Meltenyi Biotec, Inc., Auburn, CA) was added followed by a wash with PBS plus 2% FBS. Cells were then incubated for 30 min on ice with anti-CD45 (H130) (BioLegend), anti-CD3 (OKT3) (BioLegend), anti-CD8 (SK1) (BD), anti-CD56 (HCD56) (BioLegend), anti-CD38 (HIT2) (BioLegend) and live/dead fixable aqua dye (eF660, eBioscience), washed twice with PBS plus 2% FBS and then stored at 4 °C until acquired by FACS Verse (BD Biosciences, San Jose, CA). Data were analyzed using FlowJo software (Version 10.0.8, Tree Star Inc., Ashland, Or).

### Statistical analysis

Prism 7 software is used for statistical analysis. Student’s t test was performed for two-group analysis. Pearson’s correlation coefficients were calculated. *P* values less than 0.05 were considered to be statistically significant.

## Data Availability

All data referred to in the manuscript are available.

## ACKNOWLEDGEMENT

This work was supported in part by grants from Natural Science Foundation of China (NSFC, Grant No. 31991173 to CD), National key research (Grant No. 2016YFC130390 to LN) and an award from Zhejiang University Foundation (to CD).

